# Antibody responses to BNT162b2 vaccination in Japan: Monitoring vaccine efficacy by measuring IgG antibodies against the receptor binding domain of SARS-CoV-2

**DOI:** 10.1101/2021.07.19.21260728

**Authors:** Hidetsugu Fujigaki, Yasuko Yamamoto, Takenao Koseki, Sumi Banno, Tatsuya Ando, Hiroyasu Ito, Takashi Fujita, Hiroyuki Naruse, Tadayoshi Hata, Saya Moriyama, Yoshimasa Takahashi, Tadaki Suzuki, Takahiro Murakami, Yukihiro Yoshida, Yo Yagura, Takayoshi Oyamada, Masao Takemura, Masashi Kondo, Mitsunaga Iwata, Kuniaki Saito

## Abstract

**BACKGROUND:** To fight severe acute respiratory syndrome coronavirus 2 (SARS-CoV-2), which causes coronavirus disease 2019 (COVID-19), mass vaccination has begun in many countries. To investigate the usefulness of a serological assay to predict vaccine efficacy, we analyzed the levels of IgG, IgM, and IgA against the receptor binding domain (RBD) of SARS-CoV-2 in the sera from BNT162b2 vaccinated individuals in Japan.

**METHODS:** This study included 219 individuals who received two doses of BNT162b2. The levels of IgG, IgM, and IgA against RBD were measured by enzyme-linked immunosorbent assay before and after the first and second vaccination, respectively. The relationship between antibody levels and several factors including age, gender, and hypertension were analyzed. Virus-neutralizing activity in sera was measured to determine the correlation with the levels of antibodies. A chemiluminescent enzyme immunoassay (CLEIA) method to measure IgG against RBD was developed and validated for the clinical setting.

**RESULTS:** The levels of all antibody isotypes were increased after vaccination. Among them, RBD-IgG was dramatically increased after the second vaccination. The IgG levels in females were significantly higher than in males. There was a negative correlation between age and IgG levels in males. The IgG levels significantly correlated with the neutralizing activity. The CLEIA assay measuring IgG against RBD showed a reliable performance and a high correlation with neutralizing activity.

**CONCLUSIONS:** Monitoring of IgG against RBD is a powerful tool to predict the efficacy of SARS-CoV-2 vaccination and provides useful information in considering a personalized vaccination strategy for COVID-19.

## Introduction

Coronavirus disease 2019 (COVID-19) is caused by severe acute respiratory syndrome coronavirus 2 (SARS-CoV-2) infection (1). Since the initial outbreak in Wuhan, China in late 2019, a health emergency with social and economic disruptions has spread worldwide. Efforts to develop a vaccine against SARS-CoV-2 to control the global COVID-19 pandemic became a global effort and resulted in the emergency approval of several vaccines (2-6).

One of the first approved COVID-19 vaccines, BNT162b2 (Pfizer/BioNTech), has shown promising efficacy in clinical trials (7, 8). BNT162b2 is an mRNA vaccine that expresses the full prefusion spike (S) glycoprotein of SARS-CoV-2. A two-dose regimen of BNT162b2 was found to be safe with 95% efficacy in preventing symptomatic COVID-19 in persons 16 years of age or older (8). Following the authorization of COVID-19 vaccines for emergency use by the U.S. Food and Drug Administration on December 11^th^, 2020, a mass vaccination campaign began throughout the world.

Now that it has been demonstrated that the COVID-19 vaccines can induce a humoral response thereby protecting individuals from symptomatic COVID-19, several studies attempted to use serological assays to detect antibody production following COVID-19 vaccination (9-13). To monitor the efficacy of vaccines, antibody titer can be used to predict protection against SARS-CoV-2, which is already done as a routine laboratory testing for many viruses, such as measles morbillivirus, rubella virus, and hepatitis virus.

There is consensus that serological assays are helpful for predicting vaccine efficacy; however, a variety of antigens used to detect antibodies and a variety of antibody isotypes measured in these assays may cause issues with interpreting the results (14, 15). Also, to monitor the efficacy of a vaccine by measuring antibodies against SARS-CoV-2 in the clinical laboratory, it is necessary to measure antibodies that are highly correlated with the serum neutralizing activity. A previous study from our group determined the kinetics and neutralizing activity of various antigen-specific antibody isotypes against SARS-CoV-2 in the serum of patients with COVID-19 (16). This study used enzyme-linked immunosorbent assay (ELISA) to measure IgG, IgM, and IgA for various antigens, including the full-length S, S1, receptor binding domain (RBD), and nucleocapsid (N) proteins. As a result, the measurement of IgG against the RBD (RBD-IgG) not only showed good clinical performance for diagnosing SARS-CoV-2 infection, but also exhibited a high correlation with serum neutralizing activity. Therefore, monitoring RBD-IgG may be the best indicator to quantifying the immunogenicity of vaccines.

To evaluate the usefulness of measuring anti-SARS-CoV-2 antibodies to monitor vaccination efficacy, we analyzed antibody responses including RBD-IgG, -IgM, and -IgA after the first and second dose of the BNT162b2 vaccine in a well-defined cohort of employees in Japan. We also analysed how the antibody response changes in correlation with several factors such as age, gender, and hypertension. Furthermore, we examined which antibody isotypes against RBD were strongly correlated with the neutralization titer. Finally, we developed and validated a chemiluminescent enzyme immunoassay (CLEIA) method to measure RBD-IgG for clinical settings. This study not only provides valuable information with respect to antibody responses after BNT162b2 vaccination in the Japanese population, but also the usefulness of RBD-IgG for monitoring vaccine efficacy in clinical laboratories.

## Materials and Methods

### Ethics statement, participants, and sample processing

This study was approved by the Ethics Committee for Clinical Research of the Center for Research Promotion and Support at Fujita Health University (authorization number HM20-526 and HM21-167). The study was carried out with accordance to Declaration of Helsinki. All participants provided written informed consent before undergoing any study procedure.

The study included a series of 219 employees at Fujita Health University, who received two doses of BNT162b2 at Fujita Health University Hospital. The demographic and clinical characteristics of the participants are presented in Table 1. No participant had a history of SARS-CoV-2 infection. Blood samples were collected from all participants 3 times; 1) before the first vaccination, 2) an average of 14.6 days after the first vaccination, and 3) an average of 14.3 days after the second vaccination. Serum was obtained by centrifugation for 15 min at 1,500 g at room temperature, aliquoted and stored at −80°C until use.

**Table 1.** Characteristics of subjects in this cohort.

|  |  |  | Number | Age |
| --- | --- | --- | --- | --- |
| Total subjects |  |  | 219 | 45.6 ± 11.0 |
| Gender | Male |  | 69 | 47.2 ± 12.3 |
|  | Female |  | 150 | 44.8 ± 10.3 |
| Age | Male | 20-39yr | 18 | 31.6 ± 6.3 |
|  |  | 40-49yr | 18 | 43.5 ± 3.2 |
|  |  | 50-59yr | 20 | 53.8 ± 3.0 |
|  |  | > 60yr | 13 | 63.8 ± 4.3 |
|  | Female | 20-39yr | 43 | 31.6 ± 5.6 |
|  |  | 40-49yr | 55 | 45.1 ± 2.7 |
|  |  | 50-59yr | 43 | 54.2 ± 2.6 |
|  |  | > 60yr | 9 | 61.3 ± 1.7 |
| Hypertension | Male |  | 12 | 59.1 ± 5.0 |
|  | Female |  | 6 | 52.2 ± 5.3 |
| Allergies | Male |  | 26 | 43.1 ± 13.7 |
|  | Female |  | 75 | 43.0 ± 11.1 |

To compare the hypertension and control groups, we selected 45 samples (18 hypertension subjects and 27 age-matched controls) from this cohort. The subjects in the hypertension group were selected according to the following criteria: subjects diagnosed with hypertension, regardless of uses of antihypertensive medications. The subjects in the control group were selected according to the following criteria: no current medications. To compare the allergy and control groups, we selected 174 samples (101 allergy subjects and 73 age-matched controls) from this cohort. The subjects in the allergy group were selected according to the following criteria: subjects diagnosed with allergy regardless of uses of anti-allergy medications. Those in the control group were selected according to the following criteria: no current medications.

For correlation analysis between antibody titers and neutralization activity using COVID-19 patient sera, we used 53 residual serum samples from 38 patients who were admitted to Fujita Health University Hospital from 28 February 2020 to 16 September 2020.

### Measurement of RBD-IgG, IgM, and IgA by ELISA

RBD-IgG was measured using an ELISA kit from FUJIFILM Wako Pure Chemical Corporation. All procedures were performed in accordance with the manufacturer’s instructions. IgM and IgA in serum samples were measured using ELISA assays that were developed in the previous study (16). Briefly, 96-well plates (Thermo Fisher Scientific) were coated with 250 ng of recombinant RBD protein (Acro Biosystems) and used for ELISA assays. Fifty microliter of each serum sample, which were diluted 1:201 for IgA assays or 1:2010 for IgM assays in PBS containing 2% bovine serum albumin, were added per well and incubated at room temperature for 60 min. The plates were washed three times with PBS containing 0.1% Tween 20 (PBS-T), peroxidase-labelled anti-human IgM or IgA antibody (Midrand Bioproducts) was added as secondary antibody and incubated at room temperature for 60 min. After incubation, the plates were washed five times with PBS-T and substrate solution (TMB/H_2_O_2_) (FUJIFILM Wako Pure Chemical) was added and incubated at room temperature for 10 min. The reaction was stopped by the addition of 1 M HCl and the absorbance was measured at 450 nm using a 620 nm reference filter using a microplate reader (Molecular Devices). Serially diluted pooled serum from 10 patients with COVID-19 who had strong IgA positivity and 5 patients who had strong IgM positivity were used to prepare a standard curve. The OD450 of the standard of 0.015 was converted to 1 U/mL and these values were fitted onto a line graph using linear regression analysis. The antibody units in the test serum samples were then calculated from their OD450 values using the parameters estimated from the standard curve.

### SARS-CoV-2 neutralizing assay

The neutralizing assay was conducted according to the method described in a previous report with minor modifications (16, 17). Briefly, serum samples were heat-inactivated at 56°C for 30 min and then serially diluted with Dulbecco’s Modified Eagle medium (FUJIFILM Wako Pure Chemical) supplemented with 2% fetal bovine serum (Biowest), 100 U/mL penicillin and 100 mg/mL streptomycin (Thermo Fisher Scientific). The mixture of diluted sera and 100 tissue culture infectious dose 50 hCoV-19/Japan/TY-WK-521/2020 strain (EPI_ISL_408667) were incubated at 37°C and 5% CO_2_. On day 5, the plates were fixed with 20% formalin (FUJIFILM Wako Pure Chemical) and stained with crystal violet solution (Sigma-Aldrich) to evaluate the cytopathic effect. The index of the highest serum dilution factor with cytopathic effect inhibition was defined as the microneutralization test titer (MNT).

### Measurement of RBD-IgG by CLEIA

We utilized another laboratory test kit (18) for the Accuraseed automated immunoassay system (FUJIFILM Wako Pure Chemical Corporation) to develop a CLEIA method to measure RBD-IgG in serum samples. Principle and procedures of RBD-IgG measurement by Accuraceed is shown in Fig. S1. A 10 μL serum sample and 140 μL of an immune reaction buffer were added to 12.5 μg of recombinant SARS-CoV-2 RBD-bound particles and incubated at 37°C for 3 min. After the bound and free fractions were separated, 50 μL of peroxidase-labelled anti-human IgG antibody was added and incubated at 37°C for 3 min, followed by separation of the bound and free fractions. Finally, 100 μL of the substrate solution and 100 μL of the hydrogen peroxide solution were added, and the amount of light emitted per unit time was measured. Using pooled serum from the patients with COVID-19 described above in the ELISA assay, a standard curve was generated, and the sample luminescence was applied to the calibration curve to calculate the RBD-IgG antibody units in the serum samples. The basic performance of the assay such as limit of quantification, limit of detection, accuracy and precision are provided in the supplementary information (Fig. S2–S4, Table S1–S3).

### Statistical analysis

Statistical analysis was performed using GraphPad Prism version 8.4.3 for windows (GraphPad Software). Quantitative data were analyzed by a two-tailed Mann–Whitney U test and p < 0.05 was considered statistically significant. Correlation analysis was performed using Spearman’s correlation test. Linear regression was performed to assess the correlation of RBD-IgG levels between Accuraseed and ELISA.

## Results

Volunteers (n = 219) that participated in this study were vaccinated with two doses of BNT162b2 mRNA vaccine. The characteristics of the subjects are shown in Table 1. Serum was collected 3 times according to the following time points: before the first vaccination, after the first vaccination, and after the second vaccination. The antibody titer of RBD-IgG, IgM and IgA were measured by ELISA assay. All antibody titer increased after the first and second vaccinations compared with before the first vaccination, but dramatically increased after the second vaccination (Fig. 1). The IgG titers in the serum of all subjects ranged between 0.0 and 1.8 U/mL with a mean of 0.1 ± 0.2 U/mL before the first vaccination, between 0.0 and 100.9 U/mL with a mean of 11.8 ± 12.8 U/mL after the first vaccination and between 17.2 and 762.4 U/mL with a mean of 246.6 ± 153.6 U/mL after the second vaccination (Fig. 1A). The average titer after the second vaccination increased more than 20 times from the average titer after the first vaccination (p < 0.01). In serum after the second vaccination, IgM titers in all subjects ranged between 0.7 and 146.8 U/mL with a mean of 8.7 ± 13.1 U/mL and IgA titers ranged between 2.1 and 127.5 U/mL with a mean of 34.1 ± 21.5 U/mL (Fig. 1B and C).

**Figure 1.**
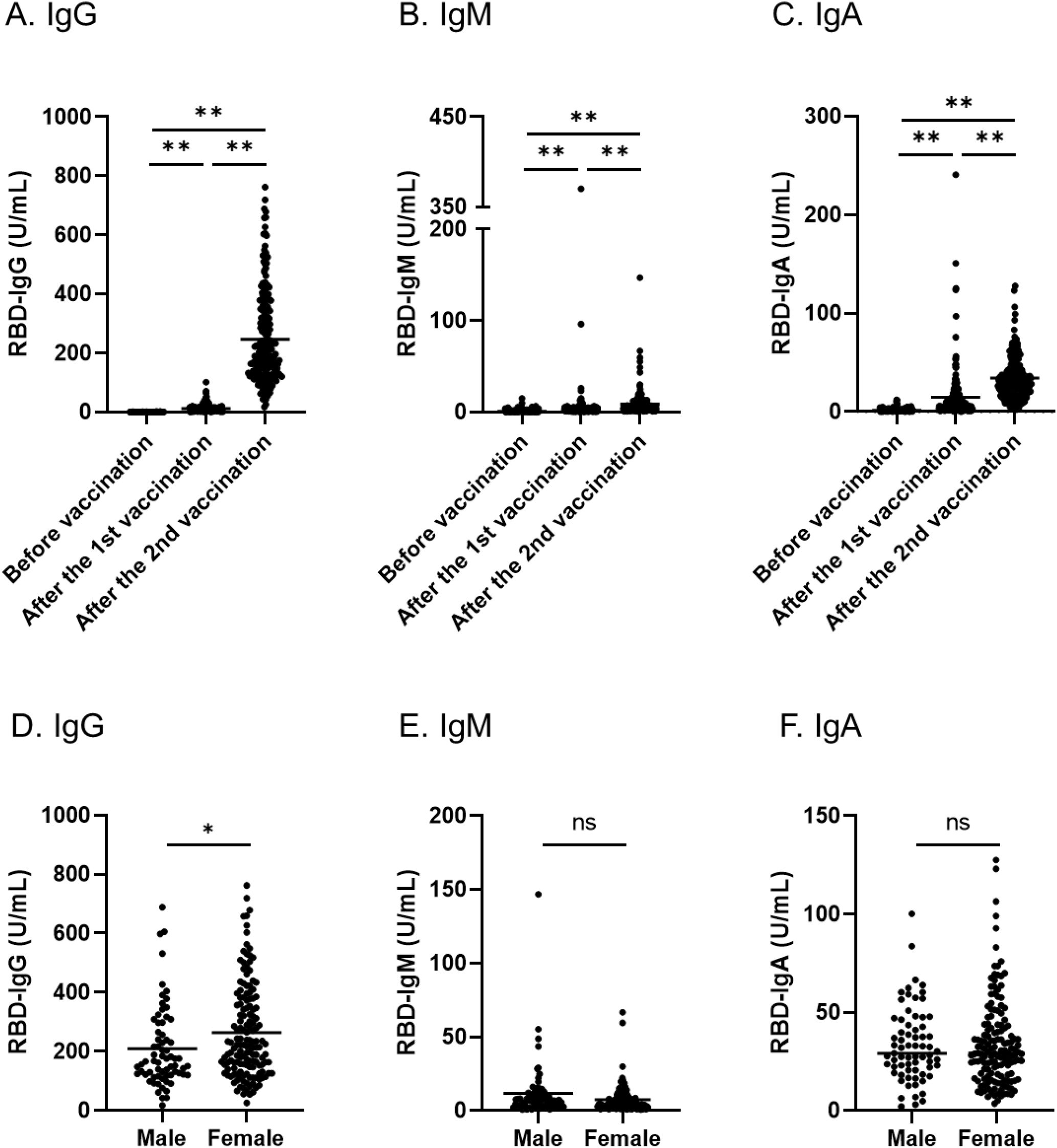
Antibody responses of IgG (A), IgM (B), and IgA (C) in all the subjects induced by BNT162b2 vaccination and differences of IgG (D), IgM (E), and IgA (F) levels between males and females after the second vaccination. Serum samples were collected before vaccination, after the first vaccination, and after the second vaccination (A-C). Antibody levels after the second vaccination were compared between males and females (D-F). The horizontal solid lines indicate the mean of antibody levels. Statistical analysis was done by a two-tailed Mann–Whitney test (*p < 0.05, **p < 0.01, ns: not significant).

The antibody titers showed differences between genders. Each antibody titter was compared between males and females. The IgG titer after the second vaccination of females was significantly higher compared with that of males (p < 0.05) (Fig. 1D). The average antibody titer was 263.6 ± 158.0 U/mL for females and was 209.9 ± 137.6 U/mL for males. Previous data showed that age was also an important factor of antibody titer after vaccination (9). Therefore, correlations were established between age and each antibody titter. In males, age correlates negatively with RBD-IgG (r = −0.410) and IgM (r = −0.283) (Fig. 2A and B, upper panel). In contrast, each antibody titter did not show a correlation with age for females (Fig. 2, lower panel). Several diseases, such as cancer, chronic kidney disease, hypertension, and diabetes, have been reported to be associated with the severity of COVID-19. The IgG titers of hypertensive subjects were significantly decreased compared to control subjects in male, but there was no difference in female (Fig. 3A). Antibody titer did not show differences between those with allergies and no medical treatment controls (Fig. 3B).

**Figure 2.**
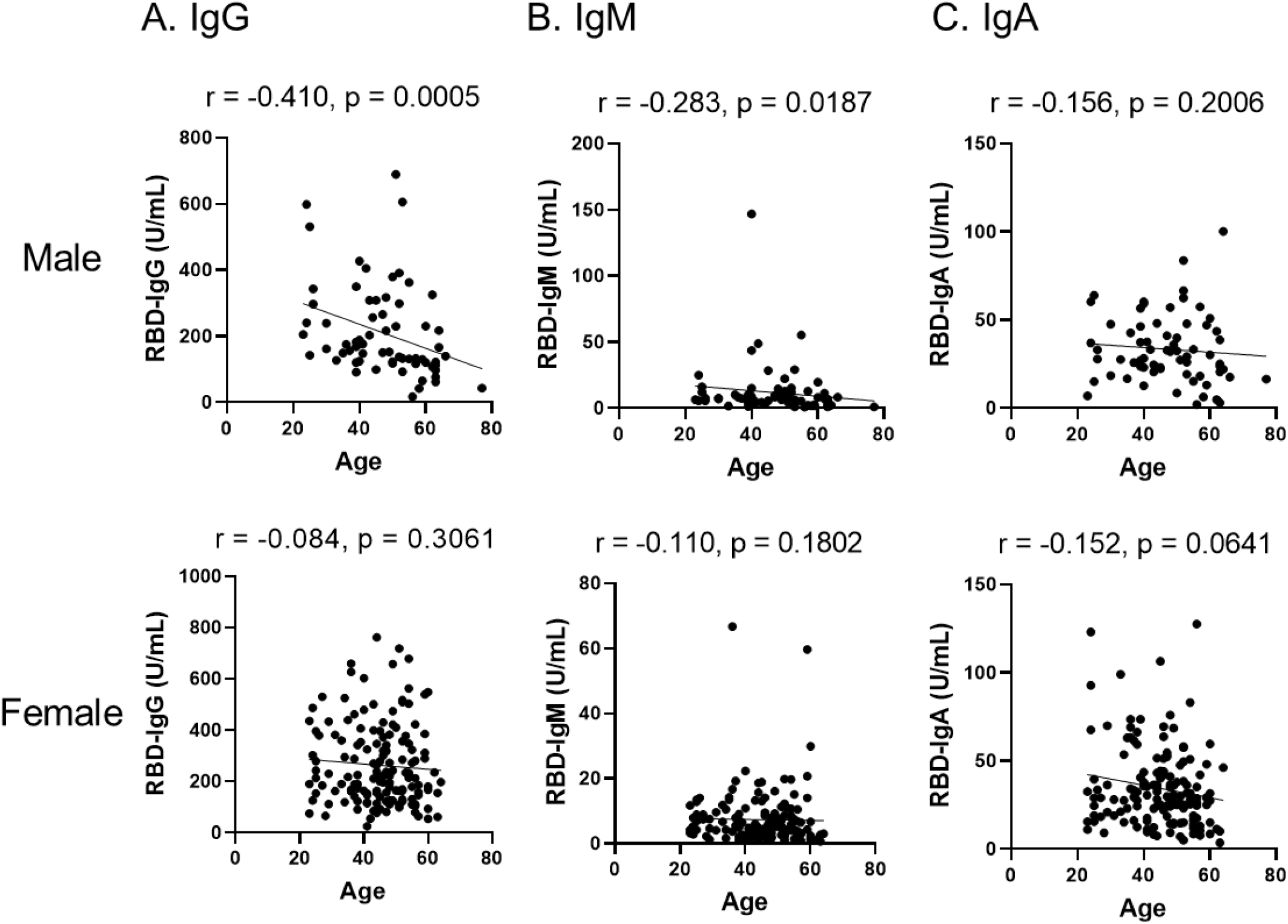
Correlation between age and IgG (A), IgM (B), and IgA (C) levels of males (upper panel) and females (lower panel) after second vaccination. Correlation analysis was calculated using a Spearman’s correlation coefficient. The values of Spearman’s r and p are presented in each figure.

**Figure 3.**
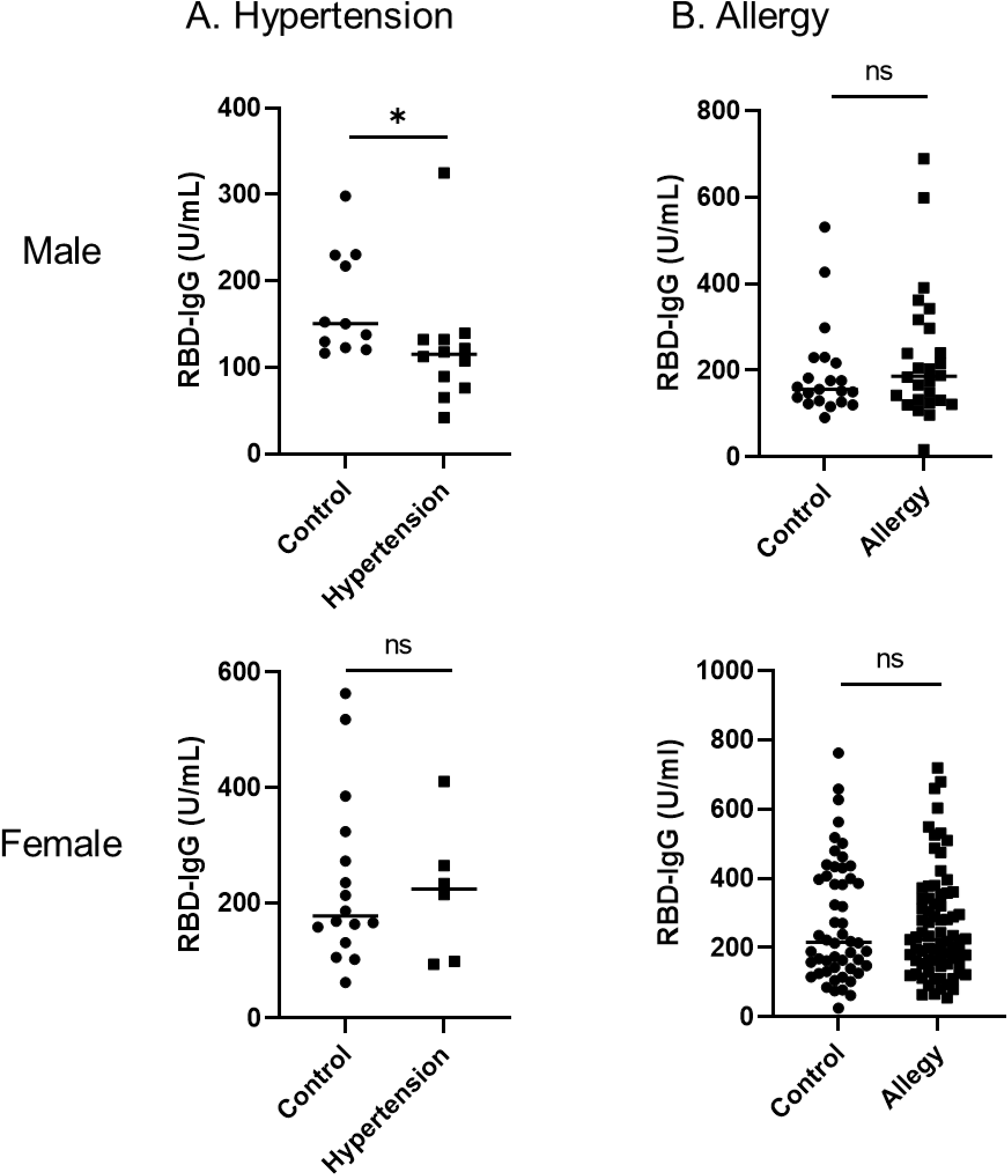
Levels of RBD-IgG of subjects who have hypertension (A) and allergy (B) (A) Subjects diagnosed with hypertension with or without antihypertensive medications (12 males and 6 females) were selected as hypertension group and compared to age-matched control group (11 males and 16 females). (B) Subjects who have allergies (26 males and 75 females) were selected as allergy group and compared to age-matched control group (21 males and 52 females). The horizontal solid lines indicate the mean of antibody levels. Statistical analysis was done by a two-tailed Mann–Whitney test (*p < 0.05, ns: not significant).

For the protection against COVID-19 infection, whether the antibody exhibits neutralizing activity produced by vaccination is the most important factor. Among 219 serum samples collected from subjects after the second vaccination, 30 samples were selected considering the distribution of antibody levels and virus-neutralizing activity was assessed. The correlation between antibody titer of IgG, IgM, and IgA and virus-neutralizing activity was examined in the serum of subjects after the second vaccination. Neutralizing activity correlated positively with IgG, IgM, and IgA titter (Fig. 4). Among the isotypes, IgG showed a higher correlation with neutralizing activity (r = 0.896).

**Figure 4.**
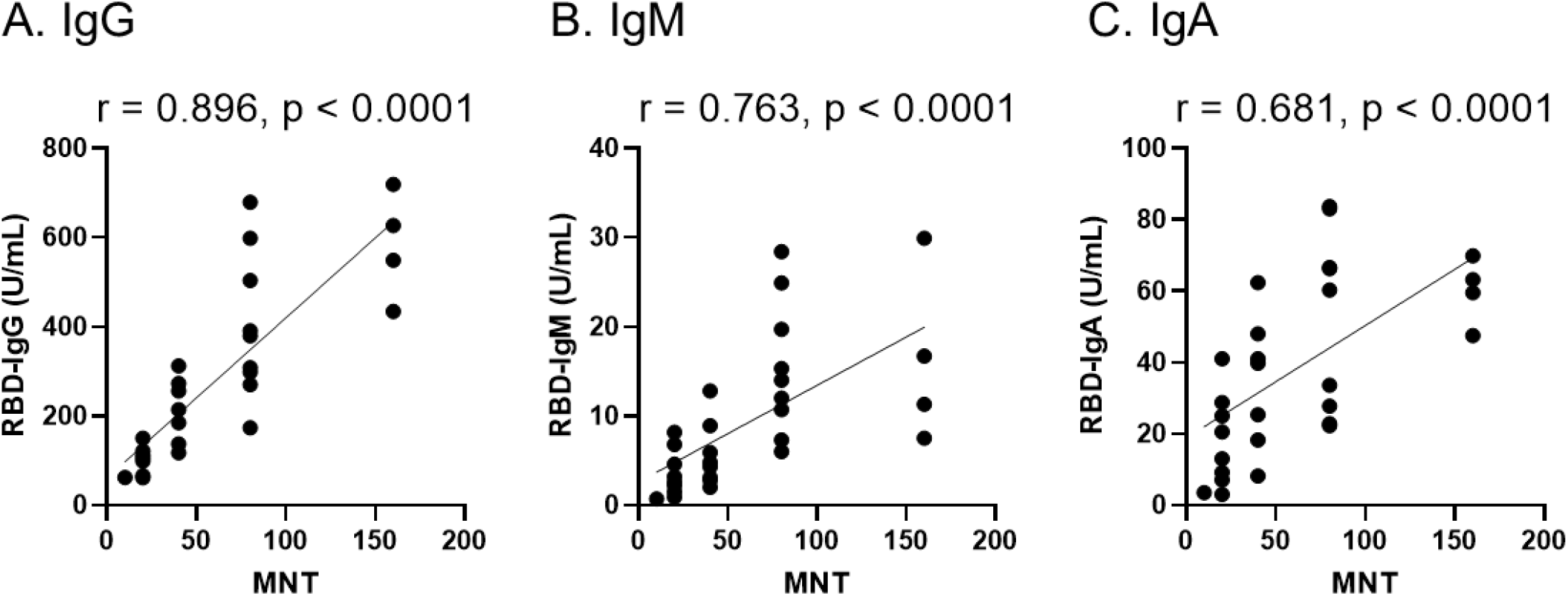
Correlation between virus neutralizing activity and IgG (A), IgM (B), and IgA (C) levels after second vaccination. Antibody measurement by ELISA and neutralization assays were performed using sera from 30 subjects after the second vaccination. The index of the highest sera dilution factor with cytopathic effect inhibition was defined as the microneutralization test titer (MNT). Correlation was calculated using Spearman’s correlation coefficient. The values of Spearman’s r and p are presented in each figure.

A rapid and accurate method to monitor antibody titers is a useful tool to investigate the efficacies of vaccination in clinical laboratories. Therefore, we developed a new CLEIA method using a two-step sandwich method to measure anti-SARS-CoV-2 antibody with the Accuraseed automated immunoassay system. This method measured antibody titers within about 10 minutes (Fig. S1). To validate the CLEIA assay, the limit of blank, limit of detection and limit of quantification, repeatability (intra-day precision) and linearity within assay range were established (Fig. S2–S4, Table S1–S3). To assess this new method, we examined the relationship between RBD-IgG levels measured by ELISA and that of the CLEIA assay. The correlation was determined between RBD-IgG levels by ELISA and RBD-IgG levels by CLEIA assay (R = 0.979, y = 0.92x-0.27) (Fig. 5A). Furthermore, the levels of the RBD-IgG by CLEIA assay correlated with the virus-neutralizing activity (r = 0.869) (Fig. 5B). Increasing evidence suggests that disease severity is positively correlated with higher antibodies, of which IgG subclasses exhibited a weakly negative correlation with viral load. We compared the RBD-IgG levels by CLEIA assay with virus-neutralizing activity in patients with COVID-19. RBD-IgG levels determined by CLEIA assay in patients with COVID-19 showed a good correlation with neutralizing activity (r = 0.723) (Fig. S5). These data indicate that the CLEIA assay is a useful tool to validate the efficiency of vaccination and monitor the severity of COVID-19 in patients.

**Figure 5.**
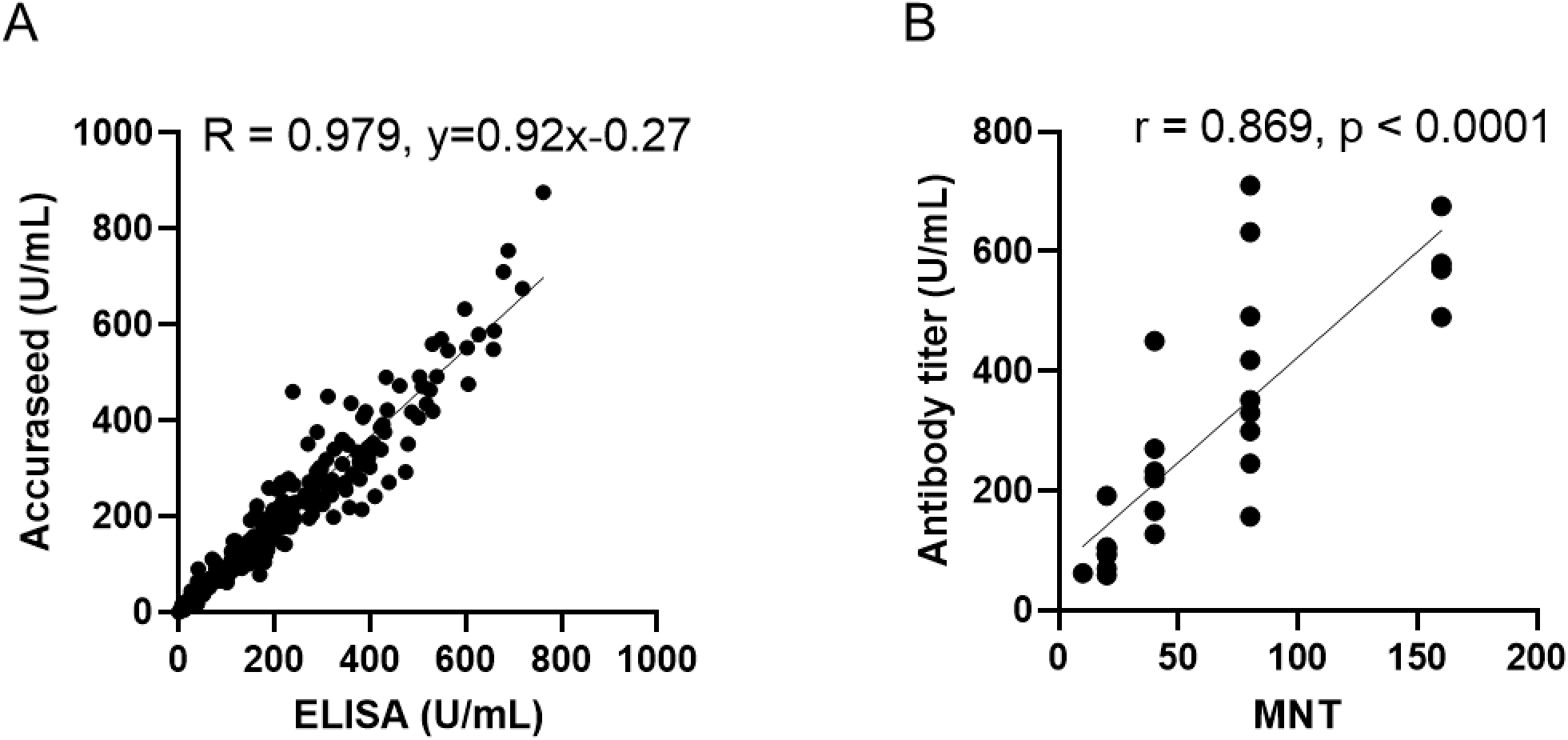
(A) Relationship between RBD-IgG levels measured by ELISA and that of the CLEIA assay. (B) Correlation between virus neutralizing activity and RBD-IgG obtained by Accuraseed. (A) RBD-IgG levels in serum samples from all subjects (total 657 samples) were measured by both ELISA and Accuraseed. Correlation R were calculated by Pearson’s correlation. (B) Antibody measurement by Accuraseed and neutralization assays were performed using sera from 30 subjects after the second vaccination. The index of the highest sera dilution factor with cytopathic effect inhibition was defined as the microneutralization test titer (MNT). Correlation was calculated using Spearman’s correlation coefficient. The values of Spearman’s r and p are presented.

## Discussion

This study investigated the antibody response produced by the SARS-CoV-2 vaccination in a well-defined cohort of employees in Japan. In all enrolled subjects, the levels of RBD-IgG, IgM and IgA antibody increased after vaccination compared to before the first vaccination. The levels of RBD-IgG in serum after the second vaccination showed a higher correlation with the virus-neutralizing activity compared with that of the other isotypes. A previous study which used COVID-19 patient serum showed that the levels of RBD-IgG were useful markers for diagnosing SARS-CoV-2 infection and neutralizing activity (16). These data indicate that the second dose vaccination potentiates neutralizing activity against COVID-19.

Persons who have been infected with SARS-CoV-2 are thought to have protective immunity and memory responses (19). IgM antibodies against SARS-CoV-2 are the first isotype to be generated against the novel antigen and can be detected as early as day 3 post COVID-19 symptoms (20). IgA antibodies also plays an important role in mucosal immunity. IgA is the most important immunoglobulin for fighting infectious pathogens in the respiratory and digestive systems at the point of pathogen entry. As an immune barrier, secretory IgA can neutralize SARS-CoV-2 before it reaches and bind to epithelial cells (20). The present data show that the antibody titter of IgG, IgM, and IgA significantly increased after the second vaccination compared with before the first vaccination (Fig. 1). In some subjects, the levels of IgA or IgM antibody increased immediately after the first vaccination and decreased after the second vaccination (Fig. 1B and C). Further studies are needed to monitor the effects of isotype differences during SARS-CoV-2 infection, but our data suggest that two doses of vaccine is valuable to protect from the infection of SARS-CoV-2 because the titer of all isotypes increased in all subjects after the second dose of vaccine.

The RBD-IgG levels after the second vaccination of males were significantly lower compared with those of females (Fig. 1D). Several studies suggest that there are many differences between males and females with respect to the immune response to SARS-CoV-2 infection and inflammatory diseases (21). Females generally produce higher levels of antibodies which remain in the circulation longer. The activation of immune cells is higher in females than males, and it correlates with the trigger of TLR7 and the production of interferon. Therefore, women with COVID-19 show a better outcome than men with the infection (22). These studies support our findings that females have a higher antibody-producing ability than males following the vaccination.

In the present study, elderly males had a lower RBD-IgG antibody-producing ability than their younger counterparts (Fig. 2A). Previous data have shown that there are age-related differences in antibody responses raised after BNT162b2 vaccination, in particular, lower frequencies of neutralizing antibodies in the elderly group (12). These data suggest that the vaccination is effective for all ages, but it is important to continue preventive measures for COVID-19, especially in elderly males at least until more longitudinal real-world data emerges.

Several diseases, such as cancer, chronic kidney disease, hypertension and diabetes, have been reported to be associated with the severity of COVID-19. The present results show that the antibody titers of hypertensive subjects in male were significantly lower compared with control subjects, but there was no statistical difference in females. Also, our analysis showed that there was no correlation between antibody titer and allergies. A previous study using a cohort of 248 health care workers in Italy reported that age and gender were statistically associated with differences in antibody response after vaccination, which is consistent with our results, whereas body mass index and hypertension had no statistically significant association. Further studies are needed to investigate what factors influence antibody responses from SARS-CoV-2 vaccination using a larger cohort.

We developed and validated a new method for measuring RBD-IgG using an automated CLEIA method. RBD-IgG measured by Accuraseed is a rapid, accurate and precise method that strongly correlates with neutralizing activity (Fig. 5, Fig. S2–S5, Tables S1–S3). Since monitoring antibody response that correlates with neutralizing activity after SARS-CoV-2 vaccination is highly advisable for predicting protection against SARS-CoV-2, this method may become a useful tool to monitor the efficacy of vaccination in clinical laboratories.

In conclusion, this study demonstrated that IgG, IgM, and IgA antibodies against RBD were produced after BNT162b2 vaccination in all subjects. Among these antibodies, RBD-IgG correlated best with neutralizing activity. These results suggest that monitoring of RBD-IgG is a powerful tool to predict efficacy of SARS-CoV-2 vaccination. The level of RBD-IgG after the second vaccination was affected by gender and age, which provide useful information to consider a personalized vaccination strategies for COVID-19.

## Data Availability

Data are available on request due to privacy or other restrictions

## Acknowledgments

We thank all the participants for participating in this study. We also thank the support from medical technologists and clinical research coordinators at the Fujita Health University Hospitals. We acknowledge Aika Kosuge, Nao Sukeda, Rika Sakai, Hitomi Kurahashi, Yumika Sugawara, Masaya Hasegawa, Honomi Ando, and Maki Namikawa for collecting the samples, and Dr. Yohei Doi for proofreading the manuscript.

## Author Contributions

H. Fujigaki, Y. Yamamoto, H. Ito, Y. Takahashi, T. Suzuki, Y. Yagura, T. Oyamada, M. Takemura, and K. Saito conceived and planned the experiments. H. Fujigaki, Y. Yamamoto, T. Ando, S. Moriyama, T. Murakami, and Y. Yoshida contributed to data acquisition and analysis. H. Fujigaki and Y. Yamamoto wrote the manuscript. T. Koseki, S. Banno, H. Ito, T. Fujita, H. Naruse, T. Hata, M. Kondo, and M. Iwata contributed to the recruitment of subjects and sample processing. H. Fujigaki, Y. Yamamoto, T. Koseki, H. Ito, Y. Takahashi, T. Suzuki, T. Murakami, Y. Yoshida, Y. Yagura, and K. Saito contributed to interpretation of the results.

## Abbreviations

SARS-CoV-2: severe acute respiratory syndrome coronavirus 2
COVID-19: coronavirus disease 2019
RBD: receptor binding domain
CLEIA: chemiluminescent enzyme immunoassay
S: spike
N: nucleocapsid
RBD-IgG: IgG against RBD
PBS-T: Phosphate buffered saline containing 0.1% Tween 20
MNT: microneutralization test titre

## References

1. Zhu N, Zhang D, Wang W, Li X, Yang B, Song J, Zhao X, et al. A Novel Coronavirus from Patients with Pneumonia in China, 2019. N Engl J Med 2020 Feb 20;382 8:727–33. Epub 2020/01/25 as doi: 10.1056/NEJMoa2001017.

2. Zhu FC, Li YH, Guan XH, Hou LH, Wang WJ, Li JX, Wu SP, et al. Safety, tolerability, and immunogenicity of a recombinant adenovirus type-5 vectored COVID-19 vaccine: a dose-escalation, open-label, non-randomised, first-in-human trial. Lancet 2020 Jun 13;395 10240:1845–54. Epub 2020/05/26 as doi: 10.1016/S0140-6736(20)31208-3.

3. Sahin U, Muik A, Derhovanessian E, Vogler I, Kranz LM, Vormehr M, Baum A, et al. COVID-19 vaccine BNT162b1 elicits human antibody and TH1 T cell responses. Nature 2020 Oct;586 7830:594–9. Epub 2020/10/01 as doi: 10.1038/s41586-020-2814-7.

4. Mulligan MJ, Lyke KE, Kitchin N, Absalon J, Gurtman A, Lockhart S, Neuzil K, et al. Phase I/II study of COVID-19 RNA vaccine BNT162b1 in adults. Nature 2020 Oct;586 7830:589–93. Epub 2020/08/14 as doi: 10.1038/s41586-020-2639-4.

5. Ewer KJ, Barrett JR, Belij-Rammerstorfer S, Sharpe H, Makinson R, Morter R, Flaxman A, et al. T cell and antibody responses induced by a single dose of ChAdOx1 nCoV-19 (AZD1222) vaccine in a phase 1/2 clinical trial. Nat Med 2021 Feb;27 2:270–8. Epub 2020/12/19 as doi: 10.1038/s41591-020-01194-5.

6. Yang J, Wang W, Chen Z, Lu S, Yang F, Bi Z, Bao L, et al. A vaccine targeting the RBD of the S protein of SARS-CoV-2 induces protective immunity. Nature 2020 Oct;586 7830:572–7. Epub 2020/07/30 as doi: 10.1038/s41586-020-2599-8.

7. Walsh EE, Frenck RW, Jr., Falsey AR, Kitchin N, Absalon J, Gurtman A, Lockhart S, et al. Safety and Immunogenicity of Two RNA-Based Covid-19 Vaccine Candidates. N Engl J Med 2020 Dec 17;383 25:2439–50. Epub 2020/10/15 as doi: 10.1056/NEJMoa2027906.

8. Polack FP, Thomas SJ, Kitchin N, Absalon J, Gurtman A, Lockhart S, Perez JL, et al. Safety and Efficacy of the BNT162b2 mRNA Covid-19 Vaccine. N Engl J Med 2020 Dec 31;383 27:2603–15. Epub 2020/12/11 as doi: 10.1056/NEJMoa2034577.

9. Pellini R, Venuti A, Pimpinelli F, Abril E, Blandino G, Campo F, Conti L, et al. Initial observations on age, gender, BMI and hypertension in antibody responses to SARS-CoV-2 BNT162b2 vaccine. EClinicalMedicine 2021 Jun;36:100928. Epub 2021/06/11 as doi: 10.1016/j.eclinm.2021.100928.

10. Padoan A, Dall’Olmo L, Rocca FD, Barbaro F, Cosma C, Basso D, Cattelan A, et al. Antibody response to first and second dose of BNT162b2 in a cohort of characterized healthcare workers. Clin Chim Acta 2021 Aug;519:60–3. Epub 2021/04/16 as doi: 10.1016/j.cca.2021.04.006.

11. Herishanu Y, Avivi I, Aharon A, Shefer G, Levi S, Bronstein Y, Morales M, et al. Efficacy of the BNT162b2 mRNA COVID-19 vaccine in patients with chronic lymphocytic leukemia. Blood 2021 Jun 10;137 23:3165–73. Epub 2021/04/17 as doi: 10.1182/blood.2021011568.

12. Muller L, Andree M, Moskorz W, Drexler I, Walotka L, Grothmann R, Ptok J, et al. Age-dependent immune response to the Biontech/Pfizer BNT162b2 COVID-19 vaccination. Clin Infect Dis 2021 Apr 27. Epub 2021/04/28 as doi: 10.1093/cid/ciab381.

13. Salvagno GL, Henry BM, Pighi L, De Nitto S, Gianfilippi GL, Lippi G. Monitoring of the immunogenic response to Pfizer BNT162b2 mRNA COVID-19 vaccination in healthcare workers with Snibe SARS-CoV-2 S-RBD IgG chemiluminescent immunoassay. Clin Chem Lab Med 2021 Jun 23. Epub 2021/06/24 as doi: 10.1515/cclm-2021-0687.

14. Lustig Y, Sapir E, Regev-Yochay G, Cohen C, Fluss R, Olmer L, Indenbaum V, et al. BNT162b2 COVID-19 vaccine and correlates of humoral immune responses and dynamics: a prospective, single-centre, longitudinal cohort study in health-care workers. Lancet Respir Med 2021 Jul 2. Epub 2021/07/06 as doi: 10.1016/S2213-2600(21)00220-4.

15. Fujigaki H, Takemura M, Osawa M, Sakurai A, Nakamoto K, Seto K, Fujita T, et al. Reliability of serological tests for COVID-19: comparison of three immunochromatography test kits for SARS-CoV-2 antibodies. Heliyon 2020 Sep;6 9:e04929. Epub 2020/09/29 as doi: 10.1016/j.heliyon.2020.e04929.

16. Fujigaki H, Inaba M, Osawa M, Moriyama S, Takahashi Y, Suzuki T, Yamase K, et al. Comparative Analysis of Antigen-Specific Anti-SARS-CoV-2 Antibody Isotypes in COVID-19 Patients. J Immunol 2021 May 15;206 10:2393–401. Epub 2021/05/05 as doi: 10.4049/jimmunol.2001369.

17. Matsuyama S, Nao N, Shirato K, Kawase M, Saito S, Takayama I, Nagata N, et al. Enhanced isolation of SARS-CoV-2 by TMPRSS2-expressing cells. Proc Natl Acad Sci U S A 2020 Mar 31;117 13:7001–3. Epub 2020/03/14 as doi: 10.1073/pnas.2002589117.

18. Ozeki Y, Tanimura Y, Nagai S, Nomura T, Kinoshita M, Shibuta K, Matsuda N, et al. Development of a New Chemiluminescent Enzyme Immunoassay Using a Two-Step Sandwich Method for Measuring Aldosterone Concentrations. Diagnostics (Basel) 2021 Mar 4;11 3. Epub 2021/04/04 as doi: 10.3390/diagnostics11030433.

19. Dan JM, Mateus J, Kato Y, Hastie KM, Yu ED, Faliti CE, Grifoni A, et al. Immunological memory to SARS-CoV-2 assessed for up to 8 months after infection. Science 2021 Feb 5;371 6529. Epub 2021/01/08 as doi: 10.1126/science.abf4063.

20. Chao YX, Rotzschke O, Tan EK. The role of IgA in COVID-19. Brain Behav Immun 2020 Jul;87:182–3. Epub 2020/05/27 as doi: 10.1016/j.bbi.2020.05.057.

21. Takahashi T, Ellingson MK, Wong P, Israelow B, Lucas C, Klein J, Silva J, et al. Sex differences in immune responses that underlie COVID-19 disease outcomes. Nature 2020 Dec;588 7837:315–20. Epub 2020/08/28 as doi: 10.1038/s41586-020-2700-3.

22. Conti P, Younes A. Coronavirus COV-19/SARS-CoV-2 affects women less than men: clinical response to viral infection. J Biol Regul Homeost Agents 2020 March-April;34 2:339–43. Epub 2020/04/08 as doi: 10.23812/Editorial-Conti-3.

