## Supplemental figures for "Antibody responses to BNT162b2 vaccination in Japan: Monitoring vaccine efficacy by measuring IgG antibodies against the receptor binding domain of SARS-CoV-2"

### Supplemental Figure S1

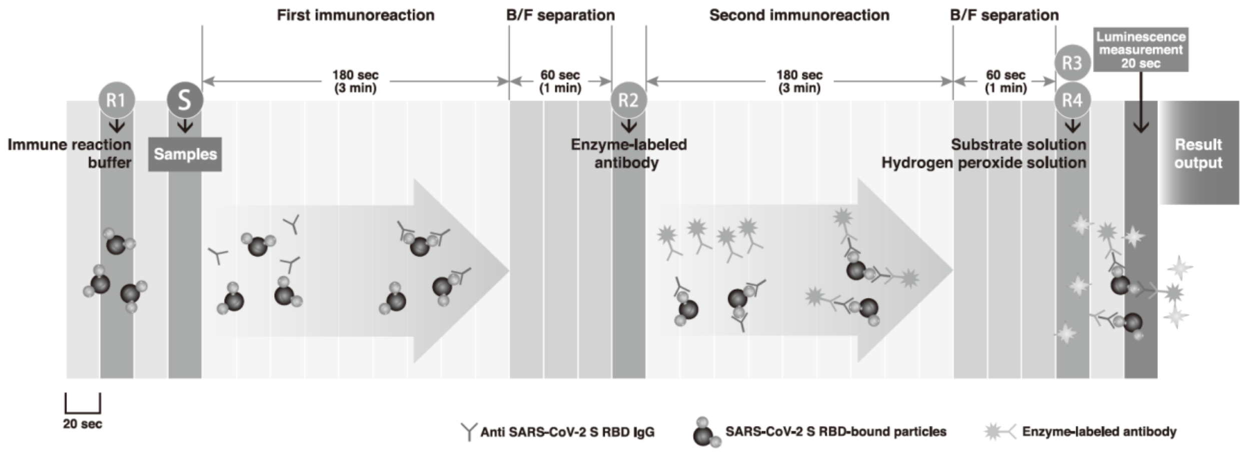


**Schematic diagram of principle and procedure of IgG measurement by Accuraceed using chemiluminescent enzyme immunoassay (CLEIA).**

**Supplemental Figure S2**

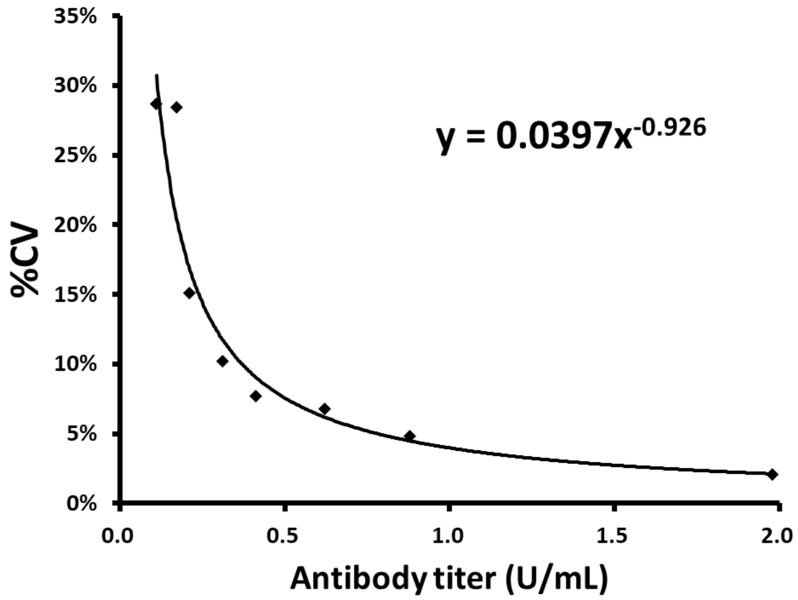


**Limit of quantitation of the antibody units using for the RBD-IgG by CLEIA assay.** The limit of blank (LoB), limit of detection (LoD) and limit of quantification (LoQ) at coefficient of variation (CV) values of 10% using the RBD-IgG by CLEIA assay were examined. These values were calculated by referring to EP17-P of the National Committee for Clinical Laboratory Standards (NCCLS).

**Supplemental Figure S3**

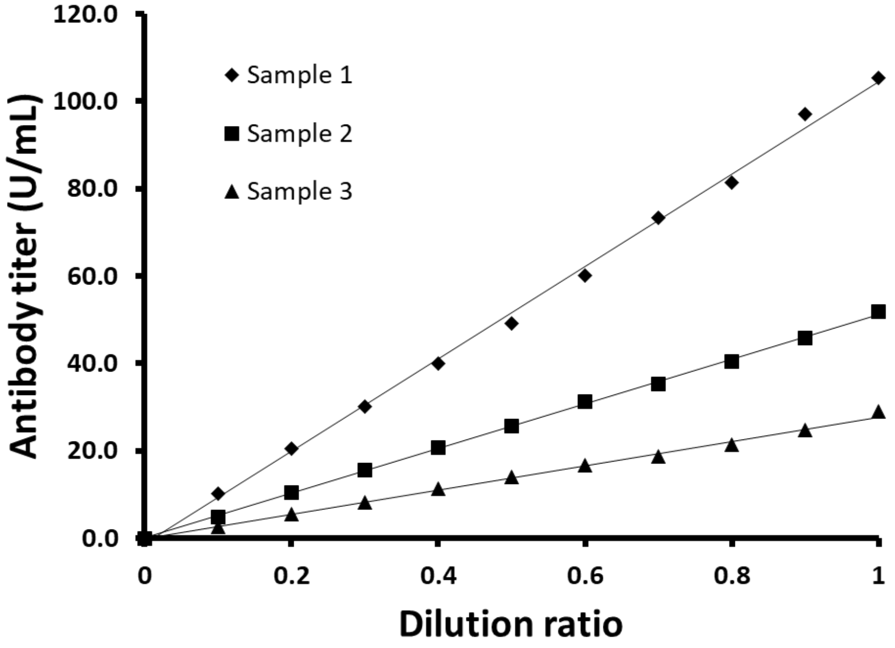


**The linearity of antibody units measured in serum samples using RBD-IgG by CLEIA assay.**

Three serum samples, from low to high concentration, were diluted 10 times with diluent solution. Linearity was established in the range from 0 to 105.3 U/mL.

**Supplemental Figure S4**

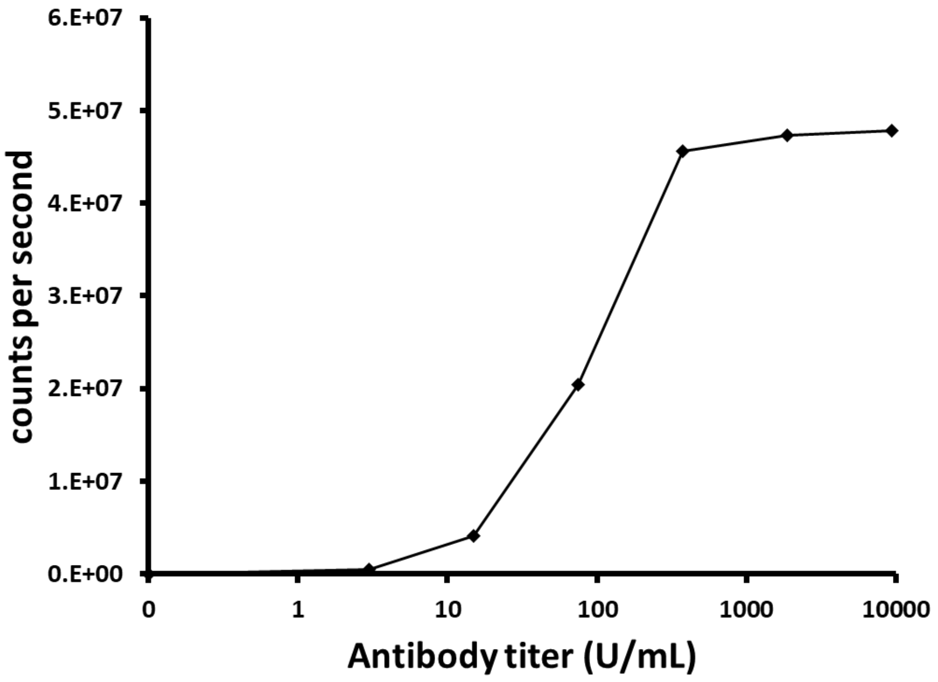


**Hook effect of antibody units using RBD-IgG by CLEIA assay.**

Antibody units were diluted from 3.0 to 9375.0 U/mL with diluent solution to evaluate the hook effect. The hook effect was not observed at concentrations up to 9375.0 U/mL.

**Supplemental Figure S5**

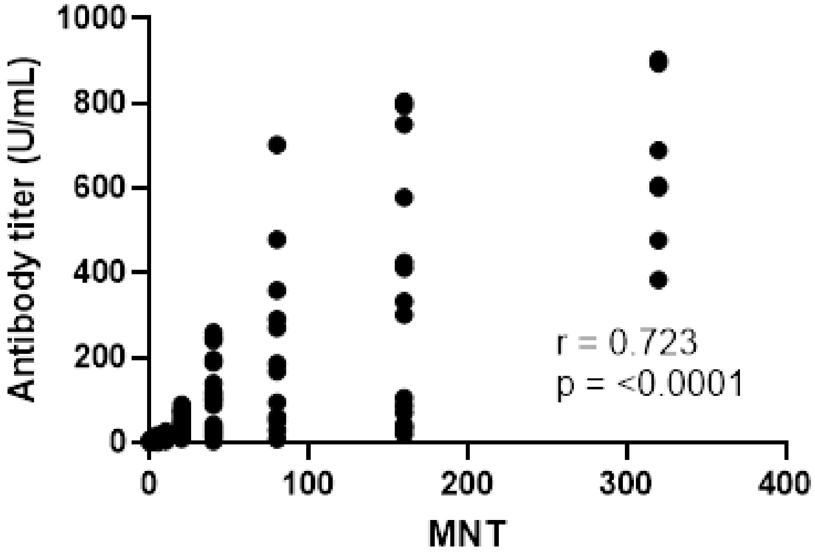


**Correlation between virus neutralizing activity and RBD-IgG levels in serum samples from patients with COVID-19.**

Antibody measurement by Accuraseed and neutralization assays were performed using 79 residual serum samples from 37 patients with COVID-19. The index of the highest sera dilution factor with cytopathic effect inhibition was defined as the microneutralization test titer (MNT). Correlation was calculated using Spearman's correlation coefficient. The values of Spearman's r and p are presented.
