## Supplemental Tables for "Antibody responses to BNT162b2 vaccination in Japan: Monitoring vaccine efficacy by measuring IgG antibodies against the receptor binding domain of SARS-CoV-2"

**Supplemental Table S1. Limit of blank (LoB), limit of detection (LoD), and limit of quantitation (LoQ) of the RBD-IgG CLEIA assay.**

| | Antibody titer (U/mL) |
|---|---|
| LoB | 0.0 |
| LoD | 0.1 |
| LoQ | 0.4 |

**Supplemental Table S2. Repeatability of the antibody measurements in serum samples using the RBD-IgG CLEIA assay. Data (mean and SD) are presented in U/mL.**

| | Sample 1 | Sample 2 | Sample 3 |
|---|---|---|---|
| N | 20 | 20 | 20 |
| Mean | 5.6 | 42.3 | 100 |
| SD | 0.1 | 1.6 | 2.7 |
| CV | 1.8% | 3.8% | 2.7% |

**Supplemental Table S3. Linearity of antibody units measured in serum samples using the RBD-IgG CLEIA assay.**

| Dilution | | Serum 1 | Serum 2 | Serum 3 |
|---|---|---|---|---|
| Antibody titer (U/mL) | 0/0 | 0.0 | 0.0 | 0.0 |
| | 1/10 | 10.1 | 4.9 | 2.7 |
| | 2/10 | 20.6 | 10.5 | 5.6 |
| | 3/10 | 30.1 | 15.5 | 8.3 |
| | 4/10 | 40.0 | 20.7 | 11.4 |
| | 5/10 | 49.1 | 25.7 | 13.9 |
| | 6/10 | 60.1 | 31.2 | 16.6 |
| | 7/10 | 73.4 | 35.2 | 18.7 |
| | 8/10 | 81.4 | 40.4 | 21.4 |
| | 9/10 | 97.0 | 45.7 | 24.7 |
| | 10/10 | 105.3 | 51.8 | 28.9 |
| Recovery rate | 1/10 | 95.9% | 94.6% | 93.4% |
| | 2/10 | 97.8% | 101.4% | 96.9% |
| | 3/10 | 95.3% | 99.7% | 95.7% |
| | 4/10 | 95.0% | 99.9% | 98.6% |
| | 5/10 | 93.3% | 99.2% | 96.2% |
| | 6/10 | 95.1% | 100.4% | 95.7% |
| | 7/10 | 99.6% | 97.1% | 92.4% |
| | 8/10 | 96.6% | 97.5% | 92.6% |
| | 9/10 | 102.4% | 98.0% | 95.0% |
| | 10/10 | 100.0% | 100.0% | 100.0% |
